# Real-world implementation and outcomes of extended-window intravenous thrombolysis in Catalonia, 2016-2024: A population-based study using the CICAT registry

**DOI:** 10.64898/2026.10.22.26366275

**Authors:** Cristina Del Valle-Vargas, Marina Martínez-Sánchez, Isabel Vielba Gomez, Yolanda Silva Blas, Antonio Doncel-Moriano, Alejandro Rodríguez-Vázquez, Blanca Lara Rodriguez, Anna Nuñez Guillen, Álvaro Garcia-Tornel, Francesc Purroy i García, Daniel Caraballo, Georgina Figueras-Aguirre, Patricia Esteve Belloch, Garbiñe Ezcurra Díaz, Ana Rodríguez-Campello, Dolores Cocho Calderón, Clara Larrañaga, Daniel Santana Moreno, Bárbara Yugueros, Xavier Jiménez, BN Salvat Mercè, Mónica Millán, Natalia Pérez de la Ossa, SCR-CAT group

## Abstract

**Background:** Evidence supporting intravenous thrombolysis (IVT) beyond 4.5 hours is based on selected populations using advanced imaging, while real-world data remains limited. We describe its uptake, selection strategies, treatment pathways, and outcomes in routine practice.

**Methods:** We conducted a population-based observational study using the prospective multicenter CICAT registry in Catalonia, Spain (2016-2024). Patients receiving extended-window IVT (>4.5 hours or unknown onset) were grouped into IVT alone and IVT before mechanical thrombectomy (bridging therapy), with conventional-window patients as a clinical reference. Excellent functional outcome was defined as a 90-day modified Rankin Scale (mRS) 0–1 or return to prestroke mRS, and good functional outcome as mRS 0–2 or return to prestroke mRS. Safety outcomes were symptomatic intracranial hemorrhage (sICH) and mortality.

**Results:** Among 12,547 patients treated with IVT, 656 (5.2%) received extended-window treatment (450 IVT alone; 206 bridging therapy). The proportion treated in the extended window increased from 3.5% in 2016-2018 to 7.4% in 2022-2024, with perfusion imaging predominantly guiding patient selection (84.1%). In the IVT alone cohort, excellent functional outcome occurred in 42.8% of extended-window and 47.2% of conventional-window patients; sICH occurred in 3.6% and 2.3%, and mortality in 12.9% and 12.5%, respectively. Corresponding outcomes in the bridging cohort 40.0% and 45.1% for good functional outcome, 20.6% and 18.6% for mortality, and 5.8% and 4.1% for sICH respectively. Differences with the reference group were also generally modest among patients aged >80 years and those with prestroke disability.

**Conclusions:** Extended-window IVT was progressively adopted across a population-based stroke network, predominantly using advanced imaging. Among selected patients, observed differences in functional and safety outcomes relative to the conventional-window reference group were generally modest, including in populations underrepresented in randomized trials. These findings should be interpreted in the context of differences in patient selection and case mix between treatment windows.

## INTRODUCTION

Intravenous thrombolysis (IVT) is a cornerstone of acute ischemic stroke (AIS) treatment. Its benefit within 4.5 hours from symptom onset is well established,^1^ with alteplase as the historical standard and tenecteplase increasingly adopted due to pharmacological and practical advantages.^2^ However, a proportion of patients present beyond this time window or with an unknown onset, such as wake-up stroke, limiting eligibility for conventional IVT. ^3^

Over the last decade, a paradigm shifts from strictly time-based to tissue-based selection has expanded the potential role of IVT beyond 4.5 hours. Trials using advanced imaging have demonstrated that selected patients treated in extended time windows can achieve improved functional outcomes, albeit with an increased risk of symptomatic intracranial hemorrhage (sICH)^4–6^ . Early evidence was provided by MRI-based mismatch studies and later reinforced by perfusion-guided trials and meta-analyses.^4–6^ Patients with large-vessel occlusion (LVO) constitute a distinct subgroup; in the extended time window, mechanical thrombectomy is the standard of care, yet the added value of IVT before thrombectomy remains uncertain. ^7–10^

Evidence from randomized trials has progressively informed routine clinical practice and guidelines. Nevertheless, real-world data on the implementation of tissue-based selection across organized stroke systems remain limited^11^. In particular, there is relatively little information on temporal patterns of adoption, imaging strategies used for patient selection and the outcomes observed in broader, unselected patient populations treated in routine practice.

Catalonia provides a suitable setting to describe this process. Acute stroke care is organized within a population-based hub-and-spoke network, and all stroke-code activations are prospectively recorded in the CICAT registry. Since 2016, extended-window IVT has been progressively incorporated with treatment decisions based on local protocols, neuroimaging findings, and treating physician judgment.

The primary aim of this study was to describe the implementation of extended-window IVT in Catalonia between 2016 and 2024, including temporal trends, hospital setting, imaging-based selection strategies, treatment pathways, and observed clinical outcomes. As a descriptive clinical reference, we report and compare clinical outcomes between patients treated in the extended and conventional time windows, separately for IVT alone and bridging therapy.

## METHODS

### Study design and population

This observational study is based on the *Codi Ictus Catalunya* (CICAT) registry, a government-mandated, prospective, hospital-based database that includes all stroke codes in Catalonia, Spain. Catalonia has a population of 7.5 million inhabitants distributed in an area of 32,000 km^2^. The Catalan Stroke network currently comprises 8 Comprehensive Stroke Centers, 6 Primary Stroke Centers and 15 Tele-Stroke Centers. The Stroke Code protocol was implemented in Catalonia in 2006 and is activated for patients with suspected acute stroke within 24 hours of symptoms onset or with unknown time of onset. The patient transport system guarantees the urgent and priority transfer of patients with suspected acute stroke to the nearest stroke center, pre-notification to the receiving center by telephone.

For the present study, we identified all consecutive patients recorded in the CICAT registry between January 2016 and December 2024 who received intravenous thrombolysis. The following variables were collected by the stroke centers participating in the CICAT registry: demographic information, pre-stroke functional status (modified Rankin Scale [mRS]), National Institutes of Health Stroke Scale (NIHSS) at admission, the presence and site of vessel occlusion, classified as proximal intracranial (terminal internal carotid artery, middle cerebral artery M1 segment, tandem occlusion, or basilar artery), distal intracranial (middle cerebral artery M2 segment, anterior cerebral artery, or posterior cerebral artery), other or unclassified (extracranial internal carotid artery, vertebral artery, or unknown occlusion site), or no occlusion; revascularization treatment (none, intravenous thrombolysis (IVT), mechanical thrombectomy (MT) with or without IVT), time metrics including: time of symptom onset or last time seen well (LTSW), stroke code activation, stroke center arrival, neuroimaging, initiation of IVT and arterial puncture. Symptomatic intracerebral hemorrhage (sICH) after revascularization treatment was defined according to the European Cooperative Acute Stroke Study II (ECASS II) criteria.^12^ Recanalization following MT was defined according to the modified thrombolysis in cerebral ischemia (mTICI) scale. ^13^ Functional outcome was centrally evaluated at 90 days (±15 days) in patients receiving reperfusion therapy through a structured telephone-based interview conducted by certified assessors using the modified Rankin scale (mRS).

To contextualize the outcomes observed among patients treated in the extended window (beyond 4.5 hours from symptom onset or with unknown time of onset), corresponding outcomes were described in patients treated within the conventional time window. Comparisons were performed separately for patients treated with IVT alone and for those treated with bridging therapy prior to MT, defined as IVT initiated within the conventional or extended window followed by MT. It is worth noting that outcomes in the conventional window group were used as a descriptive clinical reference rather than a counterfactual control group.^14^

Patient selection in the extended window was based on neuroimaging findings, including computed tomography perfusion (CTP), assessed either qualitatively (visual assessment) or quantitatively using automated software (RAPID, Syngo.via, Vitrea) and EXTEND criteria^15^ ; magnetic resonance imaging (MRI) with diffusion-weighted imaging–fluid-attenuated inversion recovery (DWI–FLAIR) mismatch following WAKE-UP criteria^16^ ; or non-contrast CT. The choice of imaging modality and treatment eligibility was based on local protocols, imaging availability and treating physician clinical judgment.

Regarding the thrombolytic agent used, all patients treated in the extended window received alteplase, as Tenecteplase was not approved for this indication in Spain during the study period. This is a real-world descriptive analysis using the population-based CICAT registry, which complies with all legal requirements mandated by the local and European law of personal data protection. In accordance with the legal procedure stablished by the Stroke Plan of the Catalan Health Department, the study was approved by the Ethics Committee at Germans Trias i Pujol Hospital (registry code PI-25-052). Informed consent was waived the present study consisted of a retrospective analysis of prospectively collected, pseudonymized registry data. This study was reported in accordance with the STROBE and RECORD guidelines^17^ . The completed RECORD checklist is provided in Supplementary Table S4.

### Study measures and clinical outcomes

The implementation of extended-window IVT was characterized in terms of temporal trends in the proportion of IVT treatments performed beyond the conventional window, hospital type, imaging-selection strategy, and treatment pathway.

Functional outcomes were described using the full distribution of the 90-day mRS and clinically relevant dichotomized measures. Excellent functional outcome was defined as an mRS score of 0–1 or return to prestroke mRS among patients treated with IVT alone, whereas good functional outcome was defined as an mRS score of 0–2 or return to prestroke mRS among patients receiving bridging therapy. Safety outcomes included sICH and 90-day mortality.

### Statistical analysis

Categorical variables were summarized as counts and percentage. Continuous and ordinal variables were described by the mean and standard deviation (SD) or median and interquartile range (IQR) as appropriate. Baseline demographic, clinical, and imaging characteristics were described according to treatment window without formal hypothesis testing.

Functional and safety outcomes were reported as observed event rates in each treatment-window group. Differences between groups were summarized using absolute differences with 95% confidence intervals, where appropriate.

The overall distribution of 90-day mRS scores was summarized using an unadjusted ordinal logistic regression model, performed separately for patients treated with IVT alone and those receiving bridging therapy. Common odds ratios (cORs) greater than 1 represented a shift toward higher mRS categories (greater disability). These analyses were intended to summarize observed differences in functional outcome distributions and not to estimate the causal effect of treatment timing.

Observed functional and safety outcomes were additionally described across prespecified clinical strata according to time from symptom onset (known onset 4.5–9 hours, known onset >9 hours, unknown onset with LTSW<9h, LTSW 9-12h and LTSW>12h) and according to selected baseline characteristics that represented exclusion criteria in prior randomized clinical trials, including pre-stroke disability (modified Rankin Scale ≥2) and age >80 years. Among patients treated in the extended window, outcomes were also described according to the neuroimaging strategy used for patient selection. These stratified analyses were exploratory and descriptive.

Analyses were based on available observations for each variable, and percentages were calculated using the corresponding available denominator. Missing data were not imputed. No formal sample-size calculation was performed because all eligible patients recorded in CICAT during the predefined study period were included. Analyses were performed using IBM SPSS Statistics, version 25.

## RESULTS

A total of 12,547 patients treated with IVT were included. Of these, 656 (5.2%) were treated in the extended time window: 450 with IVT alone and 206 as bridging therapy before mechanical thrombectomy (IVT+MT) (Figure 1). Overall, 1,227 patients were lost to 90-day follow-up, including 1,146 of 11,891 (9.6%) patients treated in the conventional time window and 81 of 656 (12.3%) patients treated in the extended time window.

**Figure 1.**
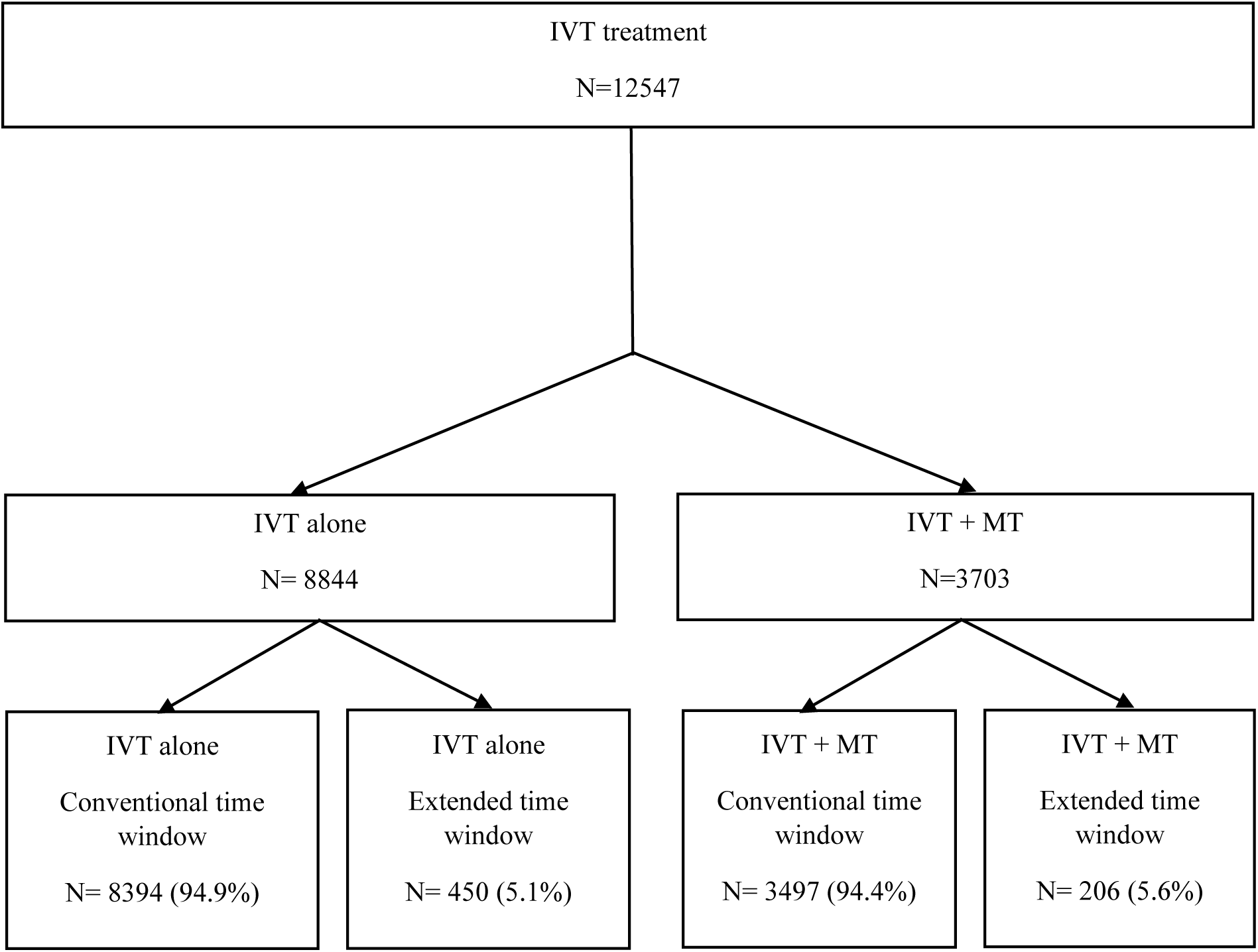
Flowchart

### Implementation of extended-window IVT

The proportion of patients treated in the extended time window increased progressively throughout the study period, raising from 3.5% in 2016–2018 to 4.8% in 2019–2021, and 7.4% in 2022–2024 (p for trend <0.001). The use of extended-window treatment also varied according to hospital type, being highest in primary stroke centers (7.0%), followed by comprehensive stroke centers (4.6%), and telestroke hospitals (1.7%) (p<0.001).

#### Patient characteristics

Baseline characteristics of patients lost to follow-up were comparable to those of patients with available 90-day outcomes, supporting the assumption that missing outcome data were largely non-differential. Table 1 summarizes the baseline characteristics of the study population.

**Table 1.**
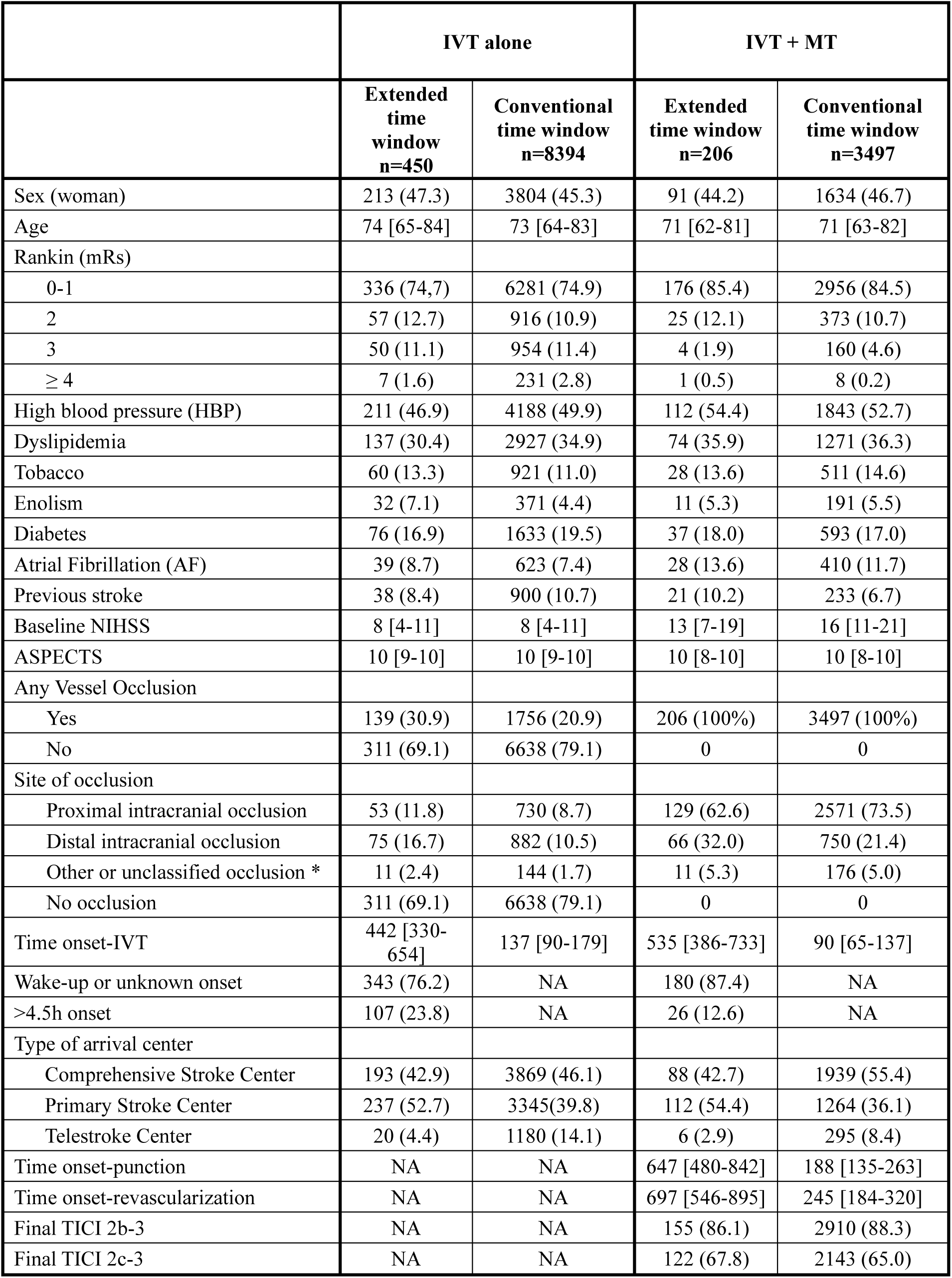
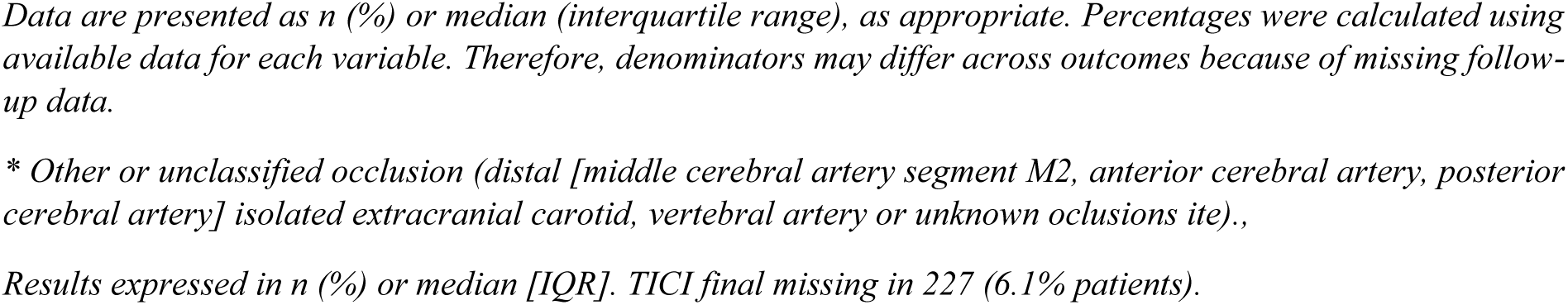
Baseline demographic, clinical and imaging characteristics of patients treated with intravenous thrombolysis alone (IVT alone) or with bridge therapy (IVT+MT), according to treatment window.

#### IVT Alone

Baseline demographics, vascular risk factors, prestroke functional status, and baseline stroke severity were similar between time windows (median NIHSS 8 [4-11] in both groups). Patients treated in the extended window had a higher prevalence of vessel occlusion (30.9% vs. 20.9%), particularly distal occlusions (16.7% vs. 10.5%) (Table S1). Median onset-to-IVT time was 442 min (IQR 330–654) in the extended window versus 137 min (IQR 90–179) in the conventional window.

#### Bridging Therapy (IVT+MT)

Extended-window patients had lower baseline NIHSS (median 13 vs. 16) and more frequently distal occlusions (32.0% vs. 21.4%). Median onset-to-IVT times was 535 vs. 90 min respectively. There were no differences in the distribution of final reperfusion grades between groups, including successful reperfusion grades (TICI 2b, 2c and 3).

### Observed functional and safety outcomes

Functional and safety outcomes according to treatment time window are shown in Table 2.

**Table 2.** Functional and safety outcomes according to treatment time window in patients treated with intravenous thrombolysis alone or with bridging therapy.

|  | IVT alone |  |  |  | IVT + MT |  |  |  |
| --- | --- | --- | --- | --- | --- | --- | --- | --- |
|  | Extended time window<br>n=450 | Conventional time window<br>n=8394 | RR<br>(95% CI) | p-value | Extended time window<br>n=206 | Conventional time window<br>n=3497 | RR<br>(95% CI) | p-value |
| Excellent functional outcome<br>(mRS 0-1 or return to previous mRS at 90 days) | 169<br>(42.8) | 3596<br>(47.2) | 0.91<br>(0.81-1.02) | 0.086 | 56<br>(31.1) | 1024<br>(32.7) | 0.95<br>(0.76-1.19) | 0.651 |
| Good functional outcome<br>(mRS 0-2 or return to previous mRS at 90 days) | 219<br>(55.4) | 4610<br>(60.5) | 0.92<br>(0.84-1.01) | 0.057 | 72<br>(40.0) | 1410<br>(45.1) | 0.89<br>(0.74-1.07) | 0.183 |
| Mortality at 90 days | 51<br>(12.9) | 954<br>(12.5) | 1.03<br>(0.79-1.34) | 0.821 | 37<br>(20.6) | 581<br>(18.6) | 1.11<br>(0.82-1.49) | 0.507 |
| sICH | 16<br>(3.6) | 196<br>(2.3) | 1.52<br>(0.92-2.51) | 0.099 | 12<br>(5.8) | 144<br>(4.1) | 1.41<br>(0.80-2.51) | 0.236 |
| Shift mRS 90 days |  |  | 1.19<br>(1.00–1.42) | 0.051 |  |  | 1.08<br>(0.83-1.40) | 0.587 |

#### IVT Alone

Among patients treated with IVT alone, excellent functional outcome at 90 days was observed in 42.8% of patients treated in the extended window and 47.2% of those treated in the conventional window. Symptomatic intracranial hemorrhage occurred in 3.6% and 2.3%, respectively, while 90-day mortality was 12.9% and 12.5%, respectively.

No statistically significant differences were observed in excellent functional outcome, sICH or 90-day mortality. The 90-day mRS distribution showed a borderline shift towards a less favorable outcome in the extended-window group, although the difference did not reach statistical significance (OR for a shift toward greater disability, 1.19; 95% CI, 1.00–1.42; p=0.051) (Figure 2).

**Figure 2.**
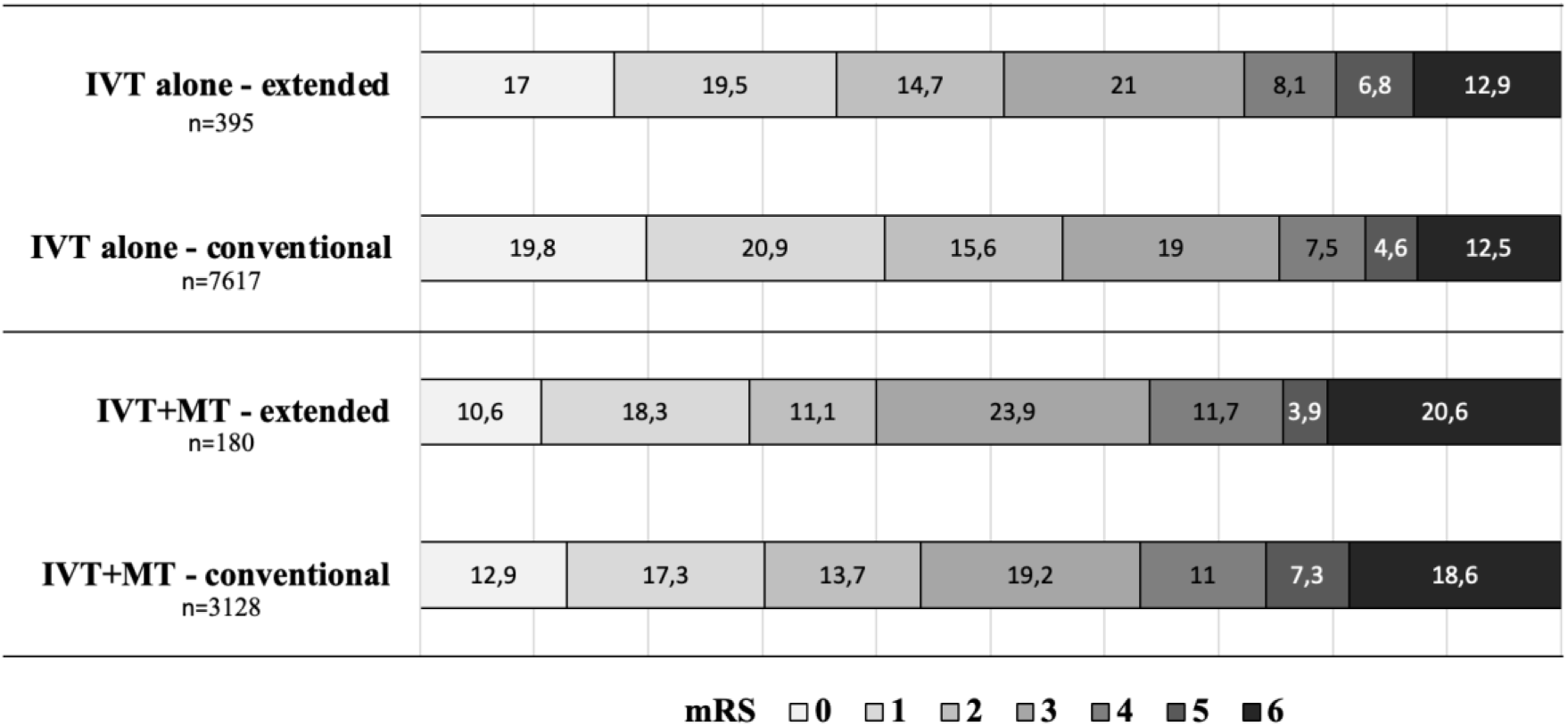
Distribution of the modified Rankin Scale at 90 days according to treatment time window in patients receiving IVT alone and IVT+MT. mRS and mortality at 90 days missing in 1227 patients (9.8%).

#### Bridging Therapy (IVT+MT)

Regarding patients treated with bridging therapy, good functional outcome at 90 days was observed in 40.0% of patients receiving IVT in the extended window and 45.1% of those treated in the conventional window. Corresponding mortality rates were 20.6% and 18.6%, and sICH occurred in 5.8% and 4.1%, respectively. No statistically significant differences were observed in good functional outcome, mortality, or sICH between treatment windows. The overall distribution of 90-day mRS scores was similar between the extended and conventional treatment windows (OR 1.08; 95% CI, 0.83–1.40; p=0.587) (Figure 2).

### Outcomes according to treatment timing and baseline characteristics

Observed outcomes across prespecified subgroups and their comparison to the conventional-time window reference group are shown in Figure 3 (good functional outcome and sICH) and Figure S1 (excellent functional outcome and 90-day mortality).

**Figure 3.**
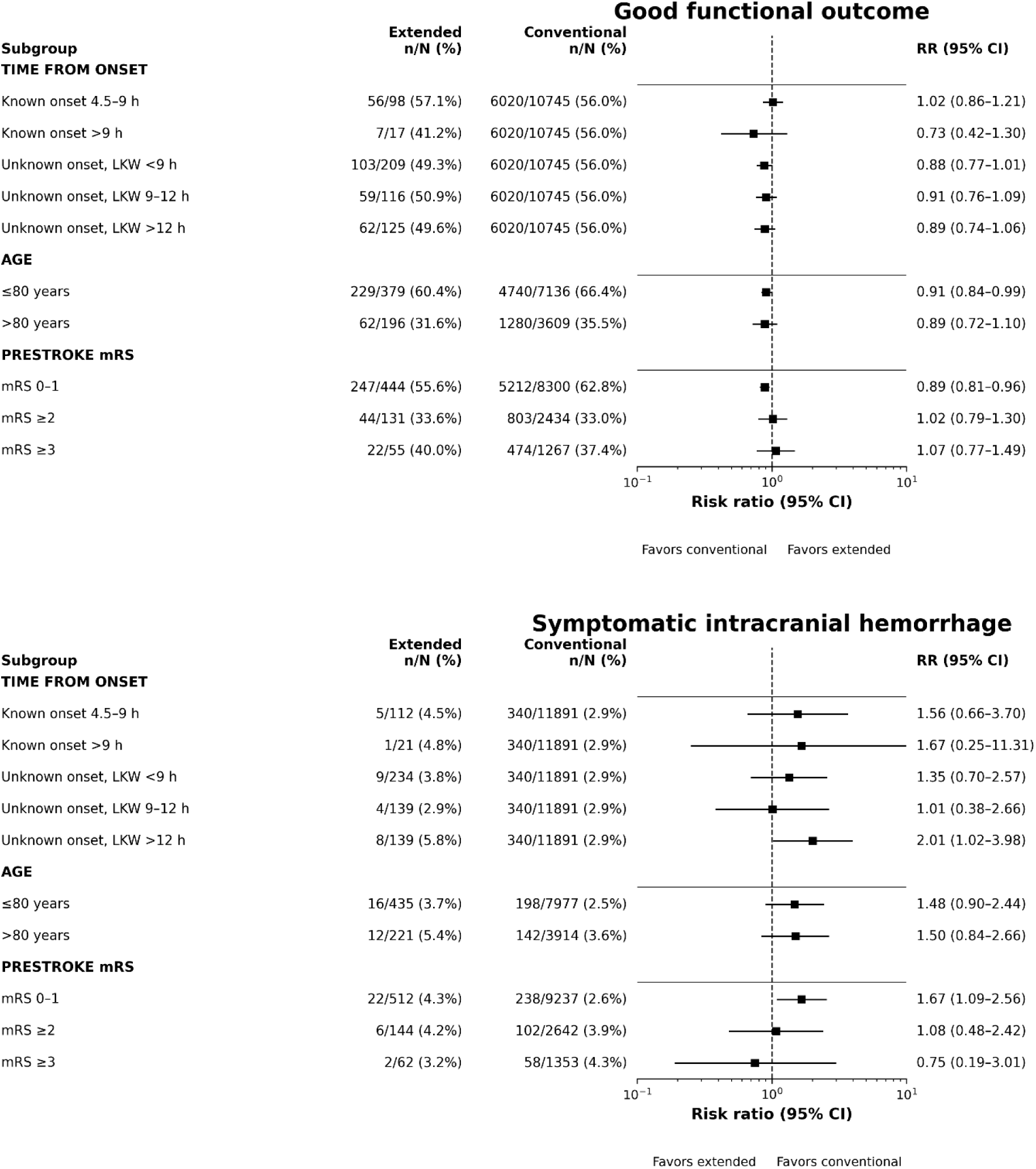
Subgroup analyses of good functional outcome and symptomatic intracranial hemorrhage according to time from onset, age and pre-stroke clinical conditions. Risk ratios are unadjusted and shown together with observed event rates. Marker size is proportional to the subgroup sample size. *Time-from-onset subgroups do not sum to the total extended-window cohort because the last-known-well time was not available in 11 patients with unknown onset (10 with available 90-day outcome*

Among patients treated in the extended time window with witnessed symptom onset, those treated between 4.5–9 hours after onset had rates of good functional outcome like those observed in the conventional-window group (57.1% vs 56.0%). In the small subgroup treated beyond 9 hours, the good functional outcome rate was lower (41.2%). Mortality was 13.3% among patients treated at 4.5–9 hours and 11.8% among those treated beyond 9 hours, comparable to the conventional-window group (14.1%).

Concerning patients with unknown symptom onset, good functional outcome was observed in 49.3%, 50.9%, and 49.6% of patients with last-known-well times of <9 hours, 9–12 hours, and >12 hours, respectively. No clear gradient in functional outcome was therefore apparent across increasing last-known-well categories. Mortality ranged from 15.3% to 16.4% across these groups. The proportion of sICH was 3.8% for LKW <9 hours, 2.9% for LKW 9–12 hours, and 5.8% for LKW >12 hours, compared with 2.9% in the reference group

For the subgroup of >80 years, good functional outcome was observed in 31.6% of those treated in the extended window and 35.5% of those treated in the conventional window. Mortality was 28.6% and 25.8% respectively, while sICH occurred in 5.4% and 3.6%.

Among patients with prestroke mRS ≥2, good functional outcome rates were 33.6% for the extended time window group and 33.0% in the conventional window group. Mortality was similar between groups (30.5% vs 29.5%), as were sICH rates (4.2% vs 3.9%).

### Imaging Modality used for extended time window selection

Information on the imaging strategy used for patient selection was available for all 656 patients treated in the extended time window. Perfusion-based imaging was the most used selection strategy, applied in 552 patients (84.1%), including qualitative visual assessment in 323 and quantitative automated software in 229. MRI DWI–FLAIR mismatch was used in 40 patients (6.1%), whereas NCCT alone was used in 64 patients (9.8%).

Observed outcomes varied across imaging-selection strategies. Patients selected using DWI–FLAIR mismatch or quantitative CT perfusion had numerically higher rates of favorable functional outcome, whereas patients selected using non-contrast CT alone had lower rates of favourable functional outcome and higher mortality (Table S2 and Figure S2).

## DISCUSSION

In this large population-based cohort, extended-window IVT was progressively incorporated into clinical practice during the study period, predominantly using advanced imaging selection. Observed differences in functional and safety outcomes between patients treated in the extended and conventional time windows were generally modest, although functional outcomes tended to be numerically less favorable in the extended-window groups. These comparisons describe outcomes among patients selected for treatment in routine clinical practice and should not be interpreted as evidence of equivalence between treatment windows or as estimates of the causal effect of treatment timing.

The temporal pattern observed in our registry closely paralleled the development of evidence supporting imaging-guided thrombolysis beyond 4.5 hours. In 2018, WAKE-UP was the first positive randomized trial to use MRI DWI-FLAIR mismatch to select patients with unknown stroke onset^16^ , followed in 2019 by EXTEND, which used perfusion imaging in patients treated 4.5-9 hours after onset or after wake-up stroke^15^ . The individual-patient-data meta-analysis by Campbell et al., also published in 2019, pooled the three major trials to the date and confirmed improved functional outcomes with alteplase in perfusion-selected patients. Excellent functional outcome occurred in 36% of alteplase-treated patients, while sICH and mortality occurred in 5% and 14%, respectively^4^ . This evidence was formally incorporated into the 2021 European Stroke Organization guidelines^18^ . In our network, extended-window IVT increased from 4.8% in 2019-2021 to 7.4% in 2022-2024, representing a relative increase of more than 50% after publication of those recommendations. Although the registry cannot attribute this change to any individual trial or guideline, the temporal association is consistent with the progressive translation of evidence into routine practice.

Since then, the evidence base has continued to expand through additional randomized trials and updated meta-analyses, further supporting the functional benefits of extended-window IVT in appropriately selected patients, despite an increased risk of symptomatic intracranial hemorrhage^6,19^. This growing body of evidence is reflected in the current AHA/ASA guidelines, which assign a Class 2a recommendation to IVT in selected patients with unknown stroke onset identified by DWI-FLAIR mismatch, and in patients who are not candidates for thrombectomy and present 4.5-9 hours after last known well, or with wake-up stroke, when salvageable tissue is demonstrated by automated perfusion imaging^20^. Nevertheless, the randomized trials underpinning these recommendations encompass heterogeneous patient populations and differ in thrombolytic agent, treatment window, imaging-selection criteria, occlusion status and use of mechanical thrombectomy (Table S3).

Within this heterogeneous evidence base, trials evaluating IVT without planned thrombectomy provide a relevant reference for our IVT alone cohort. In a recent meta-analysis by Günkan et al., including eight trials without planned thrombectomy, excellent functional outcome occurred in 43.9% of IVT-treated patients, with sICH in 2.7% and 90-day mortality in 8.4%^5^ . The observed outcomes in our cohort were broadly within the range reported in the randomized literature and in the observational TRISP cohort described by Altersberger et al., which compared conventional and extended window IVT and examined neuroimaging selection strategies, although its extended window was limited to 4.5-9 hours^11^. These comparisons are intended to provide clinical context rather than a formal cross-study comparison. Eligibility criteria, vascular characteristics, imaging selection, and outcome definitions differed across studies; notably, our definition of excellent functional outcome included return to prestroke mRS, whereas randomized meta-analyses used mRS 0-1.

Beyond clinical outcomes, the vascular characteristics of our IVT-alone cohort further illustrate the heterogeneity of patients selected for treatment in routine clinical practice. Most extended-window patients receiving IVT alone had no vessel occlusion, whereas 30.9% had an occlusion but did not undergo thrombectomy. These occlusions involved different vascular territories and were not necessarily all eligible for thrombectomy. The registry did not record the reason for not proceeding to thrombectomy in individual patients. Clinical severity, occlusion site, imaging findings, prestroke functional status, access to treatment, and changes in eligibility criteria over the nine-year study period may all have contributed to these decisions.

In contrast to IVT alone, the evidence supporting IVT before thrombectomy in the extended window remains less consistent. The TIMELESS trial did not demonstrate an improvement in 90-day functional outcome with tenecteplase in a population in which most patients subsequently underwent thrombectomy^7^. Similarly, CHABLIS-T II reported higher recanalization rates without a corresponding improvement in functional outcome^8^. In our bridging cohort, functional outcomes were numerically less favorable among patients treated in the extended window than among those treated within the conventional window, whereas the observed differences in mortality and sICH were relatively modest. Importantly, our study did not include a thrombectomy alone comparator group and therefore cannot determine the incremental benefit of IVT before thrombectomy in the extended time window.

Beyond treatment strategy, organizational pathways may also be relevant when interpreting outcomes in extended-window bridging therapy. The observational OPEN-WINDOW study, which focused on patients receiving IVT before interhospital transfer for thrombectomy, reported an association between IVT administration, higher recanalization rates during transfer, and a better three month functional outcomes^21^. In our bridging cohort, 57% of extended-window patients were initially managed at a primary stroke center or telestroke hospital and subsequently transferred for thrombectomy, while the remaining patients presented directly to a thrombectomy-capable center. These different pathways may involve variable intervals between IVT administration and arterial puncture, potentially influencing opportunities for recanalization before thrombectomy. However, our analysis was not designed to compare these pathways or determine their respective contribution to functional outcomes. The incremental benefit of IVT before thrombectomy in the extended window, particularly among patients transferred from referring hospitals, therefore remains uncertain.

Our registry also included patient groups that were absent or underrepresented in several randomized trials. As summarized in Supplementary Table S3, most trials restricted enrollment according to prestroke functional status and included relatively few patients older than 80 years. Within our cohort, patients older than 80 years had substantially lower rates of good functional outcome and higher mortality than younger patients, irrespective of treatment window. A similar pattern was observed among patients with prestroke mRS ≥2 compared with those with mRS 0-1, despite our outcome definition accounting for return to prestroke functional status. By contrast, differences in sICH according to age and prestroke disability were less pronounced. Importantly, within the subgroups of patients older than 80 years and those with prestroke disability, functional and safety outcomes were broadly similar between the extended and conventional windows. These findings highlight the importance of age and baseline functional status when interpreting clinical outcomes, although the limited subgroup sizes and selection of patients for treatment preclude conclusions regarding the effect of extended-window IVT in these populations.

Treatment timing represented another relevant dimension of clinical heterogeneity. Outcomes were similar between patients treated 4.5-9 hours after witnessed onset and those treated within the conventional window, whereas outcomes beyond 9 hours were numerically less favorable, although this subgroup was small. Among patients with unknown stroke onset, no clear gradient in outcomes according to last-known-well time was observed. These descriptive findings should be interpreted cautiously, as the influence of treatment delay cannot be disentangled from differences in vascular status, imaging selection, and patient characteristics.

Imaging selection reflected the pragmatic implementation of extended-window IVT across the stroke network. Notably, a substantial proportion of patients were selected using qualitative interpretation of CT perfusion rather than automated quantitative software. This finding suggests that implementation relied considerably on local imaging expertise and was not restricted to centers with access to automated processing, although it may also have introduced greater intercenter variability. The less favorable outcomes observed with NCCT alone should be interpreted in the context of differences in patient selection, imaging availability, local protocols, and calendar period rather than as an effect of the imaging strategy itself.

Implementation of extended-window IVT was not uniform across the stroke network, this practice was used more frequently in primary and comprehensive stroke centers and less frequently in telestroke hospitals. Differences in imaging availability, local experience, referral pathways, and access to thrombectomy may have contributed to this variation. These findings highlight the potential value of expanding access to standardized advanced imaging protocols and automated image processing in smaller centers to facilitate the implementation of guideline-recommended extended window IVT. Within a hub-and-spoke stroke network, coordinated referral pathways and telestroke services may support a more consistent adoption of these practices across the territory.

This study has several limitations. Its observational and descriptive design does not permit causal inference, and the conventional-window group should be regarded as a clinical reference rather than a counterfactual control. As CICAT is a routinely collected clinical registry not specifically designed to address the present research question, detailed information on some aspects of treatment selection and workflow was unavailable. Selection for extended-window treatment was based on local protocols, imaging availability, and physician judgment, with no central imaging adjudication, and may therefore have introduced selection bias and inter-center variability. Clinical practice evolved throughout 2016-2024, including changes in imaging availability, eligibility criteria, guideline recommendations, and reperfusion pathways. Approximately 80% of extended-window patients had an unknown time of symptom onset, so the findings predominantly reflect wake-up or unknown-onset stroke and should be extrapolated cautiously to patients with witnessed onset beyond 4.5 hours. Ninety-day follow-up was unavailable for some patients, and some prespecified subgroups were small, particularly those treated more than 9 hours after witnessed onset. Finally, all extended-window patients received alteplase, limiting generalizability to systems in which tenecteplase is now routinely used.

The study also has several strengths. CICAT prospectively captures stroke-code activity across a defined population-based network and includes patients treated at different levels of care. The nine-year observation period spans the transition from early evidence on imaging-guided thrombolysis to its broader incorporation into clinical practice. The cohort includes older patients, patients with prestroke disability, varied vascular findings and imaging-selection strategies, and both IVT-alone and bridging pathways. Functional outcome was assessed centrally at 90 days using a structured interview. Together, these features provide a broad account of how extended-window IVT has been implemented in routine stroke care and complement evidence derived from more selected randomized populations.

## CONCLUSION

In this population-based registry, extended-window IVT was progressively incorporated into routine stroke care, predominantly using advanced imaging for patient selection. Observed differences in functional and safety outcomes relative to the conventional-window reference group were generally modest among patients selected for treatment. By capturing diverse treatment pathways and a broader clinical population, including older patients and those with prestroke disability, this study provides real-world evidence on the implementation of extended-window IVT across an organized stroke network. These findings should be interpreted in the context of differences in patient selection and case mix between treatment windows.

## DATA AVAILABILITY STATEMENT

The datasets generated during and/or analyzed during the current study are available from the corresponding author on request.

## SUPPLEMENTAL MATERIAL

**Table S1.** (Supplemental). Distribution of vessel occlusion sites according to treatment strategy and time window.

|  | IVT alone |  | IVT+MT |  |
| --- | --- | --- | --- | --- |
|  | Extended time window<br>n=450 | Conventional time window<br>n=8394 | Extended time window<br>n=206 | Conventional time window<br>n=3497 |
| **TICA** | 9 (2.0) | 119 (1.4) | 10 (4.9) | 364 (10.4) |
| **MCA M1** | 40 (8.9) | 551 (6.6) | 92 (44.7) | 1647 (47.1) |
| **MCA M2** | 64 (14.2) | 674 (8.0) | 52 (25.2) | 618 (17.7) |
| **Tandem** | 0 | 9 (0.1) | 20 (9.7) | 403 (11.5) |
| **EICA** | 10 (2.2) | 105 (1.3) | 5 (2.4) | 100 (2.9) |
| **ACA** | 3 (0.7) | 56 (0.7) | 3 (1.5) | 30 (0.9) |
| **PCA** | 8 (1.8) | 152 (1.8) | 11 (5.3) | 102 (2.9) |
| **BA** | 4 (0.9) | 51(0.6) | 7 (3.4) | 157 (4.5) |
| **VA** | 1 (0.2) | 39 (0.5) | 0 | 14 (0.4) |
| **Unknown** | 0 | 0 | 6 (2.9) | 62 (1.8) |
| **No occlusion** | 311 (69.1) | 6638 (79.1) | 0 | 0 |

*Results expressed in n (%)*
*TICA: terminal internal carotid artery; MCA M1: middle cerebral artery segment M1; MCA M2: segment M2 of the middle cerebral artery; EICA: extracranial internal carotid artery; ACA: anterior cerebral artery; PCA: posterior cerebral artery; BA: basilar artery; VA: vertebral artery.*

**Table S2.** (Supplemental). Clinical characteristics and outcomes according to imaging selection strategy in patients treated with IVT in the extended time window (alone or bridge therapy).

|  | **CTP**<br>**visual**<br>**n=323** | **CTP**<br>**quantitative**<br>**n=229** | **DWI-**<br>**FLAIR**<br>**n=40** | **NCCT**<br>**n=64** |
| --- | --- | --- | --- | --- |
| **Baseline and imaging characteristics** |  |  |  |  |
| Age | 76 [64-83] | 76 [65-84] | 67 [58-76] | 76 [65-83] |
| Initial NIHSS | 8 [4-13] | 8 [5-14] | 6 [4-12] | 10 [5-18] |
| Time onset-IVT | 502<br>[343-717] | 480<br>[363-649] | 528<br>[399-558] | 455<br>[327-773] |
| Wake-up or unknown onset | 266 (82.4%) | 178 (77.7%) | 37 (92.5%) | 42 (65.6%) |
| >4.5h onset | 57 (17.6%) | 51 (22.3%) | 3 (7.5%) | 22 (34.4%) |
| Core (CVF) ml | NA | 0 [0-5] | NA | NA |
| Hipoperfusion (Tmax>6s) ml | NA | 32 [11-72] | NA | NA |
| Mismatch volume (Tmax-core) ml | NA | 26 [10-69] | NA | NA |
| Mismatch ratio | NA | 6 [3-11] | NA | NA |
| Mismatch ratio infinite (%) | NA | 20% | NA | NA |
| **Outcomes** |  |  |  |  |
| Excellent functional outcome<br>or return to pre-stroke mRS | 36.4% | 44.0% | 41.7% | 32.8% |
| Good functional outcome<br>or return to pre-stroke mRS | 47.5% | 53.5% | 63.9% | 46.6% |
| Mortality | 14.2% | 14.5% | 8.3% | 27.6% |
| sICH | 4.0% | 4.8% | 0 | 6.3% |

*Results expressed in n (%)*

**Table S3.** (Supplemental). Main inclusion criteria and outcomes of clinical trials evaluating the benefit of IVT in the extended time window.

| Trial | N | Population | Imaging | Time window | LVO (%) | MT (%) | Design | sICH risk | Clinical outcome |
| --- | --- | --- | --- | --- | --- | --- | --- | --- | --- |
| **EPITHET**<sup>22</sup><br>(2008) | 69 | ≥ 18 years<br>mRS ≤2 | MRI<br>(PWI/DWI) | 3-6h | 70 | - | tPA vs Placebo | Yes | Neutral |
| **WAKE-UP**<sup>16</sup><br>(2018) | 503 | 18-80 years<br>mRS 0-1 | MRI<br>(DWI-FLAIR) | Unknown onset | 50 | - | tPA vs Placebo | No | Benefit |
| **ECASS IV**<sup>23</sup><br>(2018) | 119 | ≥18 years<br>mRS 0-1 | MRI<br>(PWI/DWI) | 4.5-9h | 50 | 0 | tPA vs Placebo | No | Neutral |
| **EXTEND**<sup>15</sup><br>(2019) | 225 | ≥18 years<br>mRS 0-1 | CTP/<br>perfusion MRI | 4.5-9h | 70 | 0 | tPA vs Placebo | Yes | Benefit |
| **THAWS**<sup>24</sup><br>(2020) | 131 | ≥18 years<br>mRS 0-1 | MRI<br>(DWI-FLAIR) | Unknown onset | 30 | 0 | tPA 0.6 mg/kg vs Placebo | No | Neutral |
| **ROSE-TNK**<sup>25</sup><br>(2023) | 80 | ≥18 years<br>mRS 0-1 | MRI<br>(DWI-FLAIR) | 4.5-24 | NR | 0 | TNK vs Placebo | No | Neutral |
| **TWIST**<sup>26</sup><br>(2023) | 600 | ≥18 years<br>mRS 0-1 | Non-contrast CT | Unknown onset | 20 | 0 | TNK vs BMT | No | Neutral |
| **TRACE3**<sup>27</sup><br>(2024) | 516 | ≥18 years<br>mRS 0-1 | CTP | 4.5-24 | 81 | 2 | TNK vs Placebo | No | Benefit |
| **TIMELESS**<sup>7</sup><br>(2024) | 458 | ≥18 years<br>mRS ≤2 | CTP/<br>perfusion MRI | 4.5-24 | 100 | 77 | TNK + MT vs Placebo + MT | No | Neutral |
| **CHABLIS-T II**<sup>8</sup><br>(2025) | 224 | ≥18 years<br>mRS 0-1 | CTP | 4.5-24 | 100 | 55 | TNK + MT vs Placebo + MT | No | Neutral |
| **HOPE**<sup>28</sup><br>(2025) | 372 | ≥18 years<br>mRS 0-1 | CTP | 4.5-24 | 63 | 0 | tPA vs Placebo | Yes | Benefit |
| **EXPECTS**<sup>29</sup><br>(2025) | 234 | ≥18 years<br>mRS 0-1<br>Posterior circulation | Non-contrast CT | 4.5-24 | 0 | 0 | tPA vs Placebo | No | Benefit |
| **OPTION**<sup>30</sup><br>(2026) | 566 | ≥18 years<br>mRS 0-1 | CTP | 4.5-24 | 0 | 0 | TNK vs Placebo | No | Benefit |

**Table S4.** (Supplemental). The RECORD statement. Checklist of items, extended from the STROBE statement, that should be reported in observational studies using routinely collected health data.

|  | Item No. | STROBE items | Location in manuscript where items are reported | RECORD items | Location in manuscript where items are reported |
| --- | --- | --- | --- | --- | --- |
| Title and abstract |  |  |  |  |  |
|  | 1 | (a) Indicate the study's design with a commonly used term in the title or the abstract<br>(b) Provide in the abstract an informative and balanced summary of what was done and what was found | Title and Abstract | RECORD 1.1: The type of data used should be specified in the title or abstract. When possible, the name of the databases used should be included.<br>RECORD 1.2: If applicable, the geographic region and timeframe within which the study took place should be reported in the title or abstract.<br>RECORD 1.3: If linkage between databases was conducted for the study, this should be clearly stated in the title or abstract. | Title and Abstract<br>(1.1) CICAT registry<br>(1.2) Catalonia, Spain; 2016-2024<br>(1.3) Not applicable, no database linkage was performed |
| Introduction |  |  |  |  |  |
| Background rationale | 2 | Explain the scientific background and rationale for the investigation being reported | Introduction (paragraphs 2-4) |  |  |
| Objectives | 3 | State specific objectives, including any prespecified hypotheses | Introduction (final paragraph) |  |  |
| Methods |  |  |  |  |  |
| Study Design | 4 | Present key elements of study design early in the paper | Methods (study design and population) |  |  |
| Setting | 5 | Describe the setting, locations, and relevant dates, including periods of recruitment, exposure, follow-up, and data collection | Methods (study design and population) |  |  |
| Participants | 6 | (a) Cohort study - Give the eligibility criteria, and the sources and methods of selection of participants. Describe methods of follow-up<br>Case-control study - Give the eligibility criteria, and the sources and methods of case ascertainment and control selection. Give the rationale for the choice of cases and controls<br>Cross-sectional study - Give the eligibility criteria, and the sources and methods of selection of participants<br>(b) Cohort study - For matched studies, give matching criteria and number of exposed and unexposed<br>Case-control study - For matched studies, give matching criteria and the number of controls per case | Methods (study design and population) | RECORD 6.1: The methods of study population selection (such as codes or algorithms used to identify subjects) should be listed in detail. If this is not possible, an explanation should be provided.<br>RECORD 6.2: Any validation studies of the codes or algorithms used to select the population should be referenced. If validation was conducted for this study and not published elsewhere, detailed methods and results should be provided.<br>RECORD 6.3: If the study involved linkage of databases, consider use of a flow diagram or other graphical display to demonstrate the data linkage process, including the number of individuals with linked data at each stage. | ( 6.1) Methods (study design and population)<br>(6.2) + (6.3) Not applicable |
| Variables | 7 | Clearly define all outcomes, exposures, predictors, potential confounders, and effect modifiers. Give diagnostic criteria, if applicable. | Methods (study design and population, study measures and clinical outcomes) | RECORD 7.1: A complete list of codes and algorithms used to classify exposures, outcomes, confounders, and effect modifiers should be provided. If these cannot be reported, an explanation should be provided. | Not applicable, patients with stroke code activation and predefined variables were prospectively recorded directly in CICAT registry rather than derived from administrative codes. |
| Data sources/measurement | 8 | For each variable of interest, give sources of data and details of methods of assessment (measurement). Describe comparability of assessment methods if there is more than one group | Methods |  |  |
| Bias | 9 | Describe any efforts to address potential sources of bias | Methods, discussion and limitations |  |  |
| Study size | 10 | Explain how the study size was arrived at | Methods |  |  |
| Quantitative variables | 11 | Explain how quantitative variables were handled in the analyses. If applicable, describe which groupings were chosen, and why | Methods |  |  |
| Statistical methods | 12 | (a) Describe all statistical methods, including those used to control for confounding<br>(b) Describe any methods used to examine subgroups and interactions<br>(c) Explain how missing data were addressed<br>(d) Cohort study - If applicable, explain how loss to follow-up was addressed<br>Case-control study - If applicable, explain how matching of cases and controls was addressed | (a,b,c,d,e) Methods (statistical analysis) |  |  |
|  |  | Cross-sectional study - If applicable, describe analytical methods taking account of sampling strategy<br>(e) Describe any sensitivity analyses |  |  |  |
| Data access and cleaning methods |  | .. |  | RECORD 12.1: Authors should describe the extent to which the investigators had access to the database population used to create the study population.<br>RECORD 12.2: Authors should provide information on the data cleaning methods used in the study. | (12.1) The investigators had access to the complete CICAT registry population for the study period from which the study cohort was identified<br>(12.2) At each participating center, a designated collaborator reviewed the data recorded in CICAT and, when necessary, verified them against the electronic medical record or local hospital records to resolve inconsistencies or missing information. |
| Linkage |  | .. |  | RECORD 12.3: State whether the study included person-level, institutional-level, or other data linkage across two or more databases. The methods of linkage and methods of linkage quality evaluation should be provided. | Not applicable. No person-level or institutional-level linkage across separate databases was performed for the present study. |
| Results |  |  |  |  |  |
| Participants | 13 | (a) Report the numbers of individuals at each stage of the study (e.g., numbers potentially eligible, examined for eligibility, confirmed eligible, included in the study, completing follow-up, and analysed)<br>(b) Give reasons for non-participation at each stage.<br>(c) Consider use of a flow diagram | (a,b,c) Results and Figure 1. | RECORD 13.1: Describe in detail the selection of the persons included in the study (i.e., study population selection) including filtering based on data quality, data availability and linkage. The selection of included persons can be described in the text and/or by means of the study flow diagram. | Methods, First paragraph of results section and Figure 1. |
| Descriptive data | 14 | (a) Give characteristics of study participants (e.g., demographic, clinical, social) and information on exposures and potential confounders<br>(b) Indicate the number of participants with missing data for each variable of interest<br>(c) Cohort study - summarise follow-up time (e.g., average and total amount) | (a,b,c) Results, table 1. |  |  |
| Outcome data | 15 | Cohort study - Report numbers of outcome events or summary measures over time<br>Case-control study - Report numbers in each exposure category, or summary measures of exposure<br>Cross-sectional study - Report numbers of outcome events or summary measures | Results |  |  |
| Main results | 16 | (a) Give unadjusted estimates and, if applicable, confounder-adjusted estimates and their precision (e.g., 95% confidence interval). Make clear which confounders were adjusted for and why they were included<br>(b) Report category boundaries when continuous variables were categorized<br>(c) If relevant, consider translating estimates of relative risk into absolute risk for a meaningful time period | (a,c) Results (table 2 and figure 2).<br>(b) Predefined in methods (timing categories, age > 80, mRS ≥ 2)<br>(c) Absolute event rates are reported for each treatment-window group. |  |  |
| Other analyses | 17 | Report other analyses done—e.g., analyses of subgroups and interactions, and sensitivity analyses | Results |  |  |
| Discussion |  |  |  |  |  |
| Key results | 18 | Summarise key results with reference to study objectives | During the study period, there was a progressive adoption using predominantly advanced imaging. Generally, there was a modest observed differences in the extended vs conventional window. |  |  |
| Limitations | 19 | Discuss limitations of the study, taking into account sources of potential bias or imprecision. Discuss both direction and magnitude of any potential bias | Discussion | RECORD 19.1: Discuss the implications of using data that were not created or collected to answer the specific research question(s). Include discussion of misclassification bias, unmeasured confounding, missing data, and changing eligibility over time, as they pertain to the study being reported. | Discussion |
| Interpretation | 20 | Give a cautious overall interpretation of results considering objectives, limitations, multiplicity of analyses, results from similar studies, and other relevant evidence | Discussion |  |  |
| Generalisability | 21 | Discuss the generalisability (external validity) of the study results | Population-based regional hub-and-spoke network with a broad case mix. Limitations for systems using tenecteplase or different organizational pathways. |  |  |
| Other Information |  |  |  |  |  |
| Funding | 22 | Give the source of funding and the role of the funders for the present study and, if applicable, for the original study on which the present article is based | This project has been supported by grants from the Spanish Ministry of Economy and Competitiveness RICORS (Redes de Investigación Cooperativa Orientada a Resultados en Salud) de Enfermedades Vasculares Cerebrales RD24/0009/0015-0002-0006-0010-0011-0019, funded by the Instituto de Salud Carlos III. |  |  |
| Accessibility of protocol, raw data, and programming code |  | .. |  | RECORD 22.1: Authors should provide information on how to access any supplemental information such as the study protocol, raw data, or programming code. | Data available from corresponding author on reasonable request |

\*Reference: Benchimol EI, Smeeth L, Guttman A, Harron K, Moher D, Petersen I, Sørensen HT, von Elm E, Langan SM, the RECORD Working Committee. The REporting of studies Conducted using Observational Routinely-collected health Data (RECORD) Statement. *PLoS Medicine* 2015; in press.
\*Checklist is protected under Creative Commons Attribution (CC BY) license.

**Figure S1.**
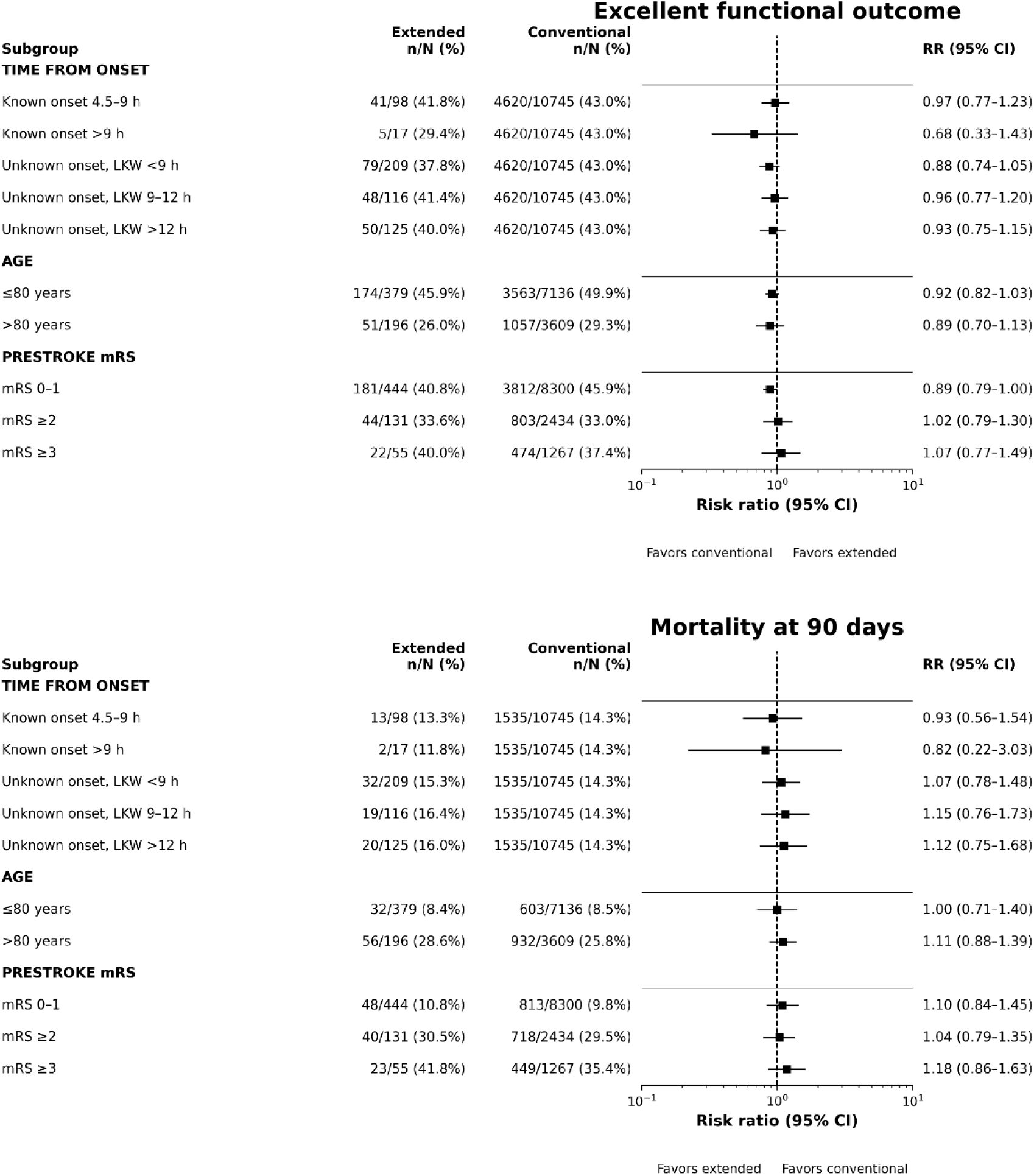
Additional subgroup analyses of excellent functional outcome and mortality according to time from onset, age and pre-stroke clinical conditions. Risk ratios are unadjusted and shown together with observed event rates. Marker size is proportional to the subgroup sample size.

**Figure S2.**
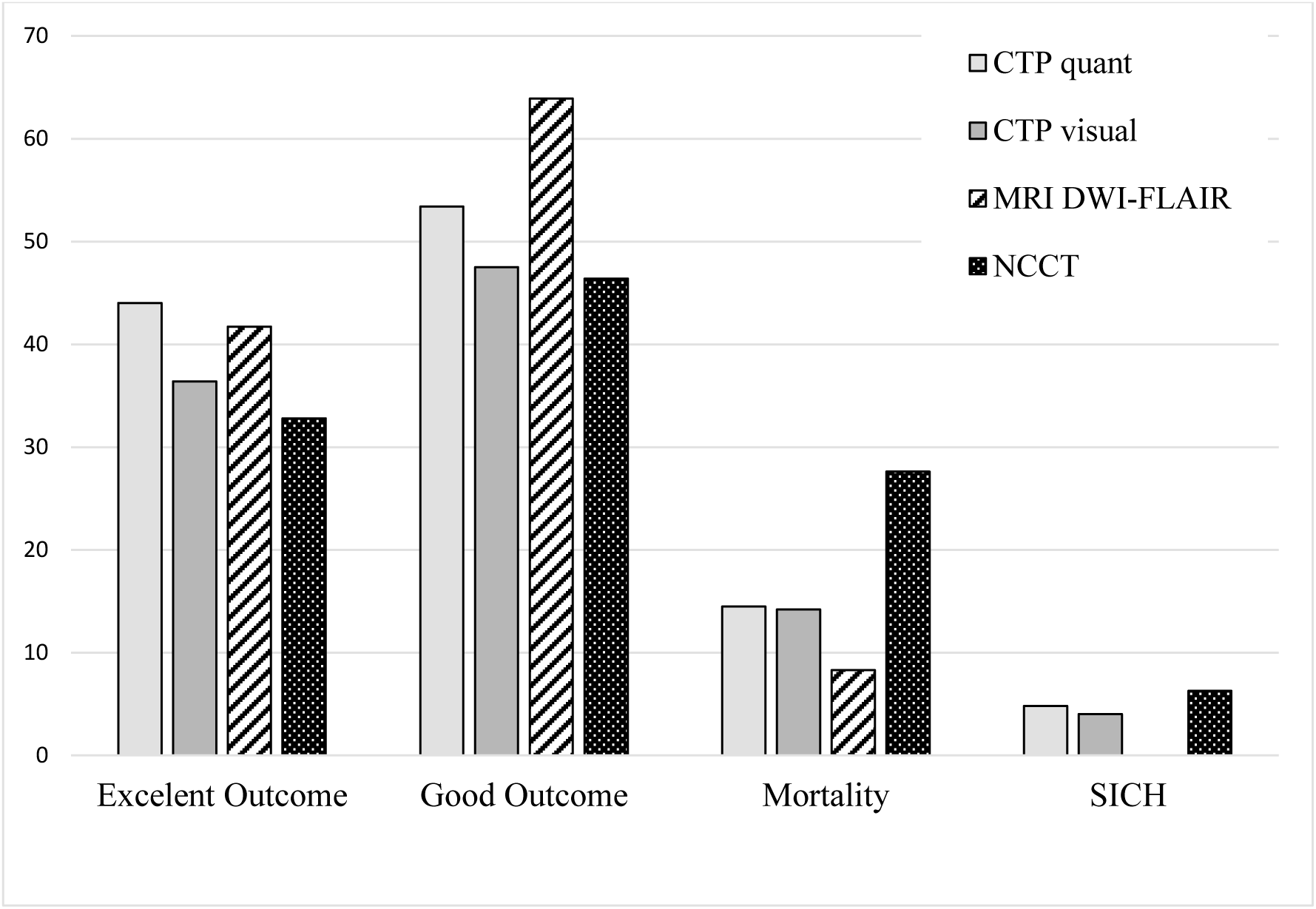
Functional and safety outcomes according to imaging selection method in patients treated with IVT alone or IVT+MT in the extended time window. CTP quant: CT perfusion with automated software quantitative assessment; CTP visual: CT perfusion without automated software quantitative assessment (eye-human evaluation); MRI DWI-FLAIR: Magnetic resonance with DWI-FLAIR mismatch criteria; NCCT: non-contrast CT.

## REFERENCES

1. Hacke W, Kaste M, Bluhmki E, Brozman M, Dávalos A, Guidetti D, Larrue V, Lees K, Medeghri Z, Machnig T, et al. Thrombolysis with alteplase 3 to 4.5 hours after acute ischemic stroke. N Engl J Med. 2008;359:1317–1329. doi:10.1056/NEJMoa0804656

2. Potla N, Ganti L. Tenecteplase vs. alteplase for acute ischemic stroke: a systematic review. Int J Emerg Med. 2022;15:1. doi: 10.1186/s12245-021-00399-w

3. Thomalla G, Gerloff C. Treatment concepts for wake-up stroke and stroke with unknown time of symptom onset. Stroke. 2015;46:2707–2713. doi: 10.1161/STROKEAHA.115.009701

4. Campbell BCV, Ma H, Ringleb PA, Parsons MW, Churilov L, Bendszus M, Levi CR, Hsu C, Kleinig TJ, Fatar M, et al. Extending thrombolysis to 4·5-9 h and wake-up stroke using perfusion imaging: a systematic review and meta-analysis of individual patient data. Lancet. 2019;394:139–147. doi: 10.1016/S0140-6736(19)31053-0

5. Günkan A, Ferreira MY, Vilardo M, Scarcia L, Bocanegra-Becerra JE, Cardoso LJC, Fabrini LF, Almeida GO, Semione G, Ferreira C, et al. Thrombolysis for ischemic stroke beyond the 4.5-hour window: A meta-analysis of randomized clinical trials. Stroke. 2025;56:580–590. doi: 10.1161/STROKEAHA.124.048536

6. Patel S, Nischal SA, Kale KM, Gooch MR, Tjoumakaris SI, Jabbour PM. Intravenous Thrombolysis Beyond the Conventional Time Window for Acute Ischemic Stroke: A Systematic Review and Meta-Analysis. JAMA Network Open. 2026;9(8):e2626990.

7. Albers GW, Jumaa M, Purdon B, Zaidi SF, Streib C, Shuaib A, Sangha N, Kim M, Froehler MT, Schwartz NE, et al. Tenecteplase for stroke at 4.5 to 24 hours with perfusion-imaging selection. N Engl J Med. 2024;390:701–711. doi: 10.1056/NEJMoa2310392

8. Cheng X, Hong L, Lin L, Churilov L, Ling Y, Yang N, Fu J, Lu G, Yue Y, Zhang J, et al. Tenecteplase thrombolysis for stroke up to 24 hours after onset with perfusion imaging selection: the CHABLIS-T II randomized clinical trial. Stroke. 2025;56:344–354. doi: 10.1161/STROKEAHA.124.048375

9. Escribano Paredes JB, Salerno A, Klug J, Dirren E, Sanda N, Bonvin C, Dunet V, Vargas M, Saliou G, Machi P, et al. Intravenous r-tPA before thrombectomy versus thrombectomy alone in strokes with unknown time of onset. Stroke. 2022;53:E136–E138. doi: 10.1161/STROKEAHA.121.037741

10. Demeestere J, Qureshi MM, Vandewalle L, Wouters A, Strbian D, Nogueira RG, Nagel S, Yamagami H, Puetz V, Abdalkader M, et al. Outcomes of bridging intravenous thrombolysis versus endovascular therapy alone in late-window acute ischemic stroke. Stroke. 2024;55:1767–1775. doi: 10.1161/STROKEAHA.124.046495

11. Altersberger VL, Sibolt G, Enz LS, Hametner C, Scheitz JF, Henon H, Bigliardi G, Strambo D, Martinez-Majander N, Stolze LJ, et al. Intravenous thrombolysis 4.5-9 Hours after stroke onset: a cohort study from the TRISP collaboration. Ann Neurol. 2023;94:309–320. doi: 10.1002/ana.26669

12. Hacke W, Kaste M, Fieschi C, Von Kummer R, Davalos A, Meier D, Larrue V, Bluhmki E, Davis S, Donnan G, et al. Randomised double-blind placebo-controlled trial of thrombolytic therapy with intravenous alteplase in acute ischaemic stroke (ECASS II). Lancet. 1998;352:1245–1251. doi: 10.1016/s0140-6736(98)08020-9

13. Tomsick T, Broderick J, Carrozella J, Khatri P, Hill M, Palesch Y, Khoury J, et al. Revascularization results in the interventional management of stroke II trial. AJNR Am J Neuroradiol. 2008;29:582–587. doi:10.3174/ajnr.A0843

14. Churilov L, Hayward K, Yogendrakumar V, Andrew N. Conducting descriptive epidemiology and causal inference studies using observational data: A 10-point primer for stroke researchers. European stroke journal. 2025;10(2):307–329.

15. Ma H, Campbell BCV, Parsons MW, Churilov L, Levi CR, Hsu C, Kleinig TJ, Wijeratne T, Curtze S, Dewey H, et al. Thrombolysis guided by perfusion imaging up to 9 hours after onset of stroke. N Engl J Med. 2019;380:1795–1803. doi: 10.1056/NEJMOA1813046

16. Thomalla G, Simonsen CZ, Boutitie F, Andersen G, Berthezene Y, Cheng B, Cheripelli B, Cho TH, Fazekas F, Fiehler J, et al. MRI-Guided thrombolysis for stroke with unknown time of onset. N Engl J Med. 2018;379:611–622. doi: 10.1056/NEJMoa1804355

17. Benchimol EI, Smeeth L, Guttmann A, Harron K, Moher D, Peteresen I, Sørensen HT, von Elm E, Langan SM. The REporting of studies Conducted using Observational Routinely-collected health Data (RECORD) statement. PLoS medicine. 2015;12(10).

18. Berge E, Whiteley W, Audebert H, Marchis GM De, Fonseca AC, Padiglioni C, Ossa NP de la, Strbian D, Tsivgoulis G, Turc G. European Stroke Organisation (ESO) guidelines on intravenous thrombolysis for acute ischaemic stroke. European stroke journal. 2021;6(1):I–LXII.

19. Pitchan Velammal PNK, Dhasakeerthi T, Manzoori D, Mummareddy SS, Duzan J, Elangovan C, Krishnaiah B. Thrombolysis in the Delayed Treatment Window for Acute Ischemic Stroke: Updated Systematic Review and Meta-Analysis. Stroke. 2026.

20. Prabhakaran S, Gonzalez NR, Zachrison KS, Adeoye O, Alexandrov AW, Ansari SA, Chapman S, Czap AL, Dumitrascu OM, Ishida K, Jadhav AP, Johnson B, Johnston KC, Khatri P, Kimberly WT, Lee VH, Leslie-Mazwi TM, Mac Grory B, Madsen TE, Menon B, Mistry EA, Park S, Parker S, Pérez de la Ossa N, Reeves M, Saiz T, Scott PA, Schwartzberg D, Sheth SA, Sporns PB, Times S, Tjoumakaris S, Wolfe SQ, Yaghi S; Peer Review Committee. 2026 Guideline for the Early Management of Patients With Acute Ischemic Stroke: A Guideline From the American Heart Association/American Stroke Association. Stroke. 2026.

21. Seners P, Nehme N, Ter Schiphorst A, Charbonneau F, Chausson N, Belley M, Wacongne A, Kaaouana O, Henry C, Girbovan A, et al. Intravenous thrombolysis use in the late time window before interhospital transfer for thrombectomy. JAMA Neurol. 2026;83:60–67. doi: 10.1001/jamaneurol.2025.4712

22. Davis SM, Donnan GA, Parsons MW, Levi C, Butcher KS, Peeters A, Barber PA, Bladin C, De Silva DA, Byrnes G, Chalk JB, Fink JN, Kimber TE, Schultz D, Hand PJ, Frayne J, Hankey G, Muir K, Gerraty R, Tress BM, Desmond PM; EPITHET investigators. Effects of alteplase beyond 3 h after stroke in the Echoplanar Imaging Thrombolytic Evaluation Trial (EPITHET): a placebo-controlled randomised trial. Lancet Neurol. 2008 Apr;7(4):299–309. doi: 10.1016/S1474-4422(08)70044-9. Epub 2008 Feb 28. PMID: 18296121.

23. Ringleb P, Bendszus M, Bluhmki E, Donnan G, Eschenfelder C, Fatar M, Kessler C, Molina C, Leys D, Muddegowda G, Poli S, Schellinger P, Schwab S, Serena J, Toni D, Wahlgren N, Hacke W; ECASS-4 study group. Extending the time window for intravenous thrombolysis in acute ischemic stroke using magnetic resonance imaging-based patient selection. Int J Stroke. 2019 Jul;14(5):483–490. Doi: 10.1177/1747493019840938. Epub 2019 Apr 4. PMID: 30947642.

24. Koga M, Yamamoto H, Inoue M, Asakura K, Aoki J, Hamasaki T, Kanzawa T, Kondo R, Ohtaki M, Itabashi R, Kamiyama K, Iwama T, Nakase T, Yakushiji Y, Igarashi S, Nagakane Y, Takizawa S, Okada Y, Doijiri R, Tsujino A, Ito Y, Ohnishi H, Inoue T, Takagi Y, Hasegawa Y, Shiokawa Y, Sakai N, Osaki M, Uesaka Y, Yoshimura S, Urabe T, Ueda T, Ihara M, Kitazono T, Sasaki M, Oita A, Yoshimura S, Fukuda-Doi M, Miwa K, Kimura K, Minematsu K, Toyoda K; THAWS Trial Investigators. Thrombolysis With Alteplase at 0.6 mg/kg for Stroke With Unknown Time of Onset: A Randomized Controlled Trial. Stroke. 2020 May;51(5):1530–1538. doi: 10.1161/STROKEAHA.119.028127. Epub 2020 Apr 6. PMID: 32248771; PMCID: PMC7185058.

25. Wang L, Dai YJ, Cui Y, Zhang H, Jiang CH, Duan YJ, Zhao Y, Feng YF, Geng SM, Zhang ZH, et al. Intravenous tenecteplase for acute ischemic stroke within 4.5-24 hours of onset (ROSE-TNK): A Phase 2, Randomized, Multicenter Study. J Stroke. 2023;25:371–377. doi: 10.5853/jos.2023.00668

26. Roaldsen MB, Eltoft A, Wilsgaard T, Christensen H, Engelter ST, Indredavik B, Jatužis D, Karelis G, Kõrv J, Lundström E, et al. Safety and efficacy of tenecteplase in patients with wake-up stroke assessed by non-contrast CT (TWIST): a multicentre, open-label, randomised controlled trial. Lancet Neurol. 2023;22:117–126. doi: 10.1016/S1474-4422(22)00484-7

27. Xiong, Y., Campbell, B. C. V., Schwamm, L. H., Meng, X., Jin, A., Parsons, M. W., Fisher, M., Jiang, Y., Che, F., Wang, L., Zhou, L., Dai, H., Liu, X., Pan, Y., Duan, C., Xu, Y., Xu, A., Zong, L., Tan, Z., Ye, W., … TRACE-III Investigators (2024). Tenecteplase for Ischemic Stroke at 4.5 to 24 Hours without Thrombectomy. The New England journal of medicine, 391(3), 203–212. 10.1056/NEJMoa2402980

28. Zhou Y, He Y, Campbell BCV, Liebeskind DS, Yuan C, Chen H, Zhang Y, Yi T, Luo Z, Zhang Z, Meng C, Cheng J, Ouyang H, Hu J, Wang F, Zhang S, Fang Q, Hu H, Zhang X, Chen Y, Zhong W, Lansberg MG, Yan S, Lou M; HOPE investigators. Alteplase for Acute Ischemic Stroke at 4.5 to 24 Hours: The HOPE Randomized Clinical Trial. JAMA. 2025 Sep 2;334(9):788–797. doi: 10.1001/jama.2025.12063. PMID: 40773205; PMCID: PMC12332759.

29. Yan S, Zhou Y, Lansberg MG, Liebeskind DS, Yuan C, Yu H, Chen F, Chen H, Zhang B, Mao L, et al. Alteplase for posterior circulation ischemic stroke at 4.5 to 24 Hours. N Engl J Med. 2025;392:1288–1296. doi: 10.1056/NEJMoa2413344

30. Ma G, Mo R, Zuo Y, Ma Q, Zhao G, Yao X, Liang J, Zhou L, He Y, Long F, et al. Tenecteplase for acute non–large vessel occlusion 4.5 to 24 hours after ischemic stroke: the OPTION randomized clinical trial. JAMA. 2026;335:1137–1147. doi: 10.1001/jama.2026.0210

